# Variety of Energy-Dense Foods Does Not Affect Body Mass Index but Influences Food Quantity: Evidence from a Cross-Sectional Study among Adolescents in Mumbai, India

**DOI:** 10.1101/2023.07.27.23293289

**Authors:** Elina Dawoodani, Chanda Gokhale

## Abstract

**Introduction:** Over the past three decades, adolescents’ share of caloric consumption from foods that are energy-dense but low in micronutrients has increased dramatically. Additionally, the rise in overweight and obesity in this age group is one of the most significant public health challenges of recent times.

**Objective:** We intended to investigate the relationship between consuming a variety of energy-dense foods and body mass index in adolescents.

**Research Methods and Procedures:** In this analytical cross-sectional study, 373 adolescents between the age of 13-15 years old were randomly recruited from three private schools in Mumbai, India. Data on their weekly consumption of energy-dense foods, anthropometric measurements, along with other critical information was obtained using a survey and food frequency questionnaire. Categories of energy-dense foods considered included chat (Indian savory snack) and street foods, appetizers, main course, continental foods, aerated drinks, baked foods, Indian sweets, farsan (fried Indian snack), and packaged foods.

**Results:** Amongst the participants (n =373, mean age 13.4 years), 39.7% were underweight, 46.1% had a normal BMI, 9.6% were overweight, and 4.5% were obese. They reported consuming an average of 4 varieties and 10.6 servings of energy-dense foods a day. Though the consumption of a variety of energy-dense foods was not found to be associated with body mass index, it was found to be significantly, positively, and strongly associated with the number of servings consumed, indicating that variety does increase total food consumption.

**Conclusion:** In this cohort of adolescents, overall consumption of a variety of energy-dense foods does not seem to influence body mass index. Longitudinal studies are warranted to assess the impact of variety on total body composition.

## Introduction

Obesity is the most serious dietary issue afflicting children and adolescents across the world. Obese children and adolescents suffer from a variety of medical and psychological health complications, such as hypertension, impaired pulmonary function, sleep apnea, gallstone formation, insulin resistance, diabetes, low self-esteem, and eating disorders (1). In India alone, the prevalence of overweight and obesity (OW/O) has increased from 9.7% in 2001 to 19.3% in 2010 (2), and now to 23.9% in 2021 (3), indicating the need for population-based preventative measures to ensure that adolescents are not afflicted by chronic health problems at a young age.

Among the myriad elements that influence OW/O, two components have been identified to have a robust effect on food consumption and weight. First, dietary variety, which when in excess causes hyperphagia, and is associated with the development and maintenance of obesity. (4–11). Animal and human studies have shown that food consumption increases when there is more variety in a meal or diet and that greater dietary variety is associated with increased body weight and fat. (12) It is generally believed that sensory-specific satiety is the mechanism by which variety influences food intake. (11,13–15).

The second component that contributes substantially to OW/O is the consumption of energy-dense foods (EDF), which often results in energy imbalance, wherein energy intake is greater than energy expenditure. This is especially relevant in countries like India, where due to nutrition transition, which was caused by a complex amalgam of marketing, social, and economic activities, has significantly altered the lifestyles and eating patterns, especially of urban Indians (16,17). Traditional home-cooked meals have been displaced in urban Indian households by high-energy, ready-to-eat, processed foods as a result of the rapid emergence of multinational fast food chains in the country’s food sector and the influence of Western culture (17,18). Studies have indicated that the consumption of energy-dense and nutrient-poor foods and sugar-sweetened beverages has increased substantially, particularly, in Indian urban regions due to food globalization (19–21). Energy-dense foods, by definition, are those that provide a high number of calories per gram or serving while offering low amounts of essential nutrients such as vitamins, minerals, and fiber. According to the World Cancer Research Fund/American Institute for Cancer Research, the cut-off value for dietary energy density was determined at 900 kJ/100 g (215 KCal/100 g) (22), equivalent to a high energy density, and based on a recently published paper, foods such as sweets, chocolate, cake, cookies and biscuits, sweet and salty snacks, sugar-sweetened and artificially sweetened drinks, and alcoholic drinks are classified as energy dense nutrient-poor food and drinks (23). Younger generations are more influenced by new foods particularly when these are introduced through an advertising campaign that targets the group specifically (24). A recent Indian study has also reported that the risk factors which showed a statistically significant association with adolescent obesity were increased intake of fast food, increased intake of chocolates/sweets, inadequate intake of fruits, and freedom given to adolescents in buying snacks without any restriction by parents (25). Similar findings are also reported in an earlier study, where dietary pattern characterized by increased intake of carbonated soft drinks, junk food (like pizza, samosa), sweets/ candies/chewing gums/chocolates, potato chips/popcorns/packed food is associated with increased risk of these of overweight and obesity among adolescents (26).

Although there are multiple pieces of evidence of both dietary variety and consumption of EDF individually, there are inconsistencies on how these two together affect the prevalence of OW/O in adolescents globally. Moreover, to our knowledge, there are no studies that have assessed the relationship between the consumption of variety of energy-dense foods (VEDF) and the prevalence of OW/O specifically in Indian adolescents. Therefore, the primary goal of the present study was to identify the relationship between VEDF and BMI among adolescents between the age of 13-15 years belonging to the higher socioeconomic strata in Mumbai, India.

The secondary objectives of the study were as follows:

1. To quantify the consumption of VEDF and the number of servings of EDF in this population
2. To study the association of the number of servings of energy-dense foods with the BMI of adolescents.
3. To identify the prevalence of OW/O in the population using anthropometric measurements.
4. To identify other factors that would, with or without VEDF, increase BMI in adolescents.

## Methodology

### Participants

The study was limited to healthy adolescents ranging from 13 to 15 years old, belonging to high socio-economic strata, and providing written consent from the parents or caregivers to participate in the study. Students who attended private schools were selected since they are generally expected to enjoy good health due to better health awareness and lack of economic constraints. This cross-sectional study was conducted on 373-school going adolescents living in Mumbai, India, using the personal interview method of the food frequency questionnaire. Schools were randomly selected from three different Municipal wards in Mumbai. One school was selected from wards L, H/E, and H each.

### Sample Size Estimations

Before the main study commenced, a pilot study of 30 children was undertaken, and this data was analyzed for reliability. The reliability of the data was computed using Cronbach’s alpha, which shows a value of 0.707. This explains 70.7% of the total variance in the variables included in the analysis. The general rule is that α of 0.6-0.7 indicates an acceptable level of reliability, and 0.8 or greater is a good level (27). Once the reliability of the pilot study was established, the sample size was estimated based on the census of the total adolescent population in Mumbai, with a 95% confidence interval and margin of error of 0.5, bringing the total necessary sample size to 384.

### Data Collection

Data collection was conducted between 3^rd^ January and 14^th^ April 2015 using the personal interview method. The survey consisted of general information such as age, gender, occupation, food preferences, etc. The participants’ recall was used to gather information on physical activity, including the type, duration, and frequency, as well as sleeping schedules. Since our goal was to understand the effect of VEDF on BMI, variables such as type and level of activity, sleep, etc. were our potential confounders.

To understand the consumption of EDF, we identified 206 commonly consumed foods and drinks that are usually high in fat, added sugars, and sodium, contributing to the energy density of the meal while containing low levels of important micro-nutrients (28). Using a food frequency questionnaire, these 206 foods were categorized into 9 different groups, viz; (1) chats (Indian savory snack) and street foods, (2) main course, (3) appetizers, (4) aerated drinks and juices, (5) continental foods, (6) desserts and baked foods, (7) Indian sweets, (8) farsan (fried Indian snack), and (9) packaged snack items, based on their nutritional profiles and culinary categorization. We recorded the frequency over the period of the last seven days and portion sizes of all these aforementioned foods. There are various valid time frames for recall, such as the past 3 or 7 days and in the case of some foods, the past month (29). All these questions were asked in private to prevent the participants from sharing incorrect information due to peer pressure or embarrassment. To estimate the serving size of each EDF consumed by adolescents in our study, we used a picture-based method as there are no standardized serving sizes for all foods, particularly for traditional dishes or home-cooked meals. We presented participants with pictures of plates, bowls, and other commonly used containers, and prompted them to report the quantity of each food item they consumed. We then estimated their serving sizes based on their reported quantities and our visual references.

### Anthropometrics

Participants’ weight and height were measured using calibrated instruments. Autocal Solutions Private Limited, Mumbai, calibrated the weighing scale (Titanshah 2003A) and measuring tape prior to the data collection. BMI was calculated as weight in kilograms per height in meters squared. BMI z-score (zBMI) was calculated using the Centers for Disease Control and Prevention (CDC) modified growth reference standards (38).

### Ethics

The study tools and research methodologies were subjected to standard ethical approval. All the procedures used in this study were reviewed and approved in writing by the Inter System Biomedica Ethics Committee (ISBEC), Vile Parle (West), Mumbai. Prior to data collection, the principals of the respective schools were approached. After acquiring permission from the authorities, the students were addressed and given a small booklet that had both a consent letter and volunteer information sheet. This sheet provided them with all the necessary information related to the project. The students were asked to take the booklet home so that the information could be passed on to the parents too. If the child and the parent/guardian were willing to participate, informed written consent was taken. We ensured that the confidentiality of participants was maintained throughout the study.

### Statistical Analysis

Data entry and coding were done using Microsoft Excel 2010 and analysis was conducted using the R package (Version 4.1.3). Data points that did not fall within three SD of the mean were identified as outliers and removed (30). Kolmogorov-Smirnov non-parametric tests were then carried out to measure the goodness of fit and establish that the sample had a normal distribution. Due to the large sample size, the sampling distribution was considered to be approximately normal and robust for parametric testing using the central limit theorem (31). Marginal distributions were examined to identify the prevalence of underweight, normal weight, and OW/O in the given population. Independent t-tests were used to examine if mean VEDF differed significantly by gender and age. Spearman’s correlations were then carried out to examine whether there is a significant association between the response variable, zBMI, and predictor variables, such as VEDF, %VEDF, number of servings of EDF, % variety of each food group, number of time and duration of exercise, and duration of sleep. Regression analyses and linear-by-linear associations were done to test if there is a causal relationship between zBMI and VEDF, %VEDF, number of servings, income, sleep, and each of the food groups individually. Furthermore, confounders were identified using correlation matrices, added to the regression models, and re-run. The variance inflation factor was calculated to gauge if the variance of VEDF is influenced, or inflated, by its interaction/correlation with the other independent variables. Additionally, Somers’ delta was used to assess the relationship between ordinal values of VEDF and BMI. Lastly, odds ratios were estimated to predict the odds of developing OW/O due to the consumption of VEDF and the number of serving of EDF. Statistical significance for all analyses was set at p < 0.05.

## Results and Interpretations

### Population Characteristics

A total of 373 school-going adolescents were recruited into the study. Amongst the participants, 56.8% (n=212) were between 13-14 years old, 62.19% (n=232) were girls, and most of the participants were non-vegetarians (89.5%, n=334).

**Table 1.**
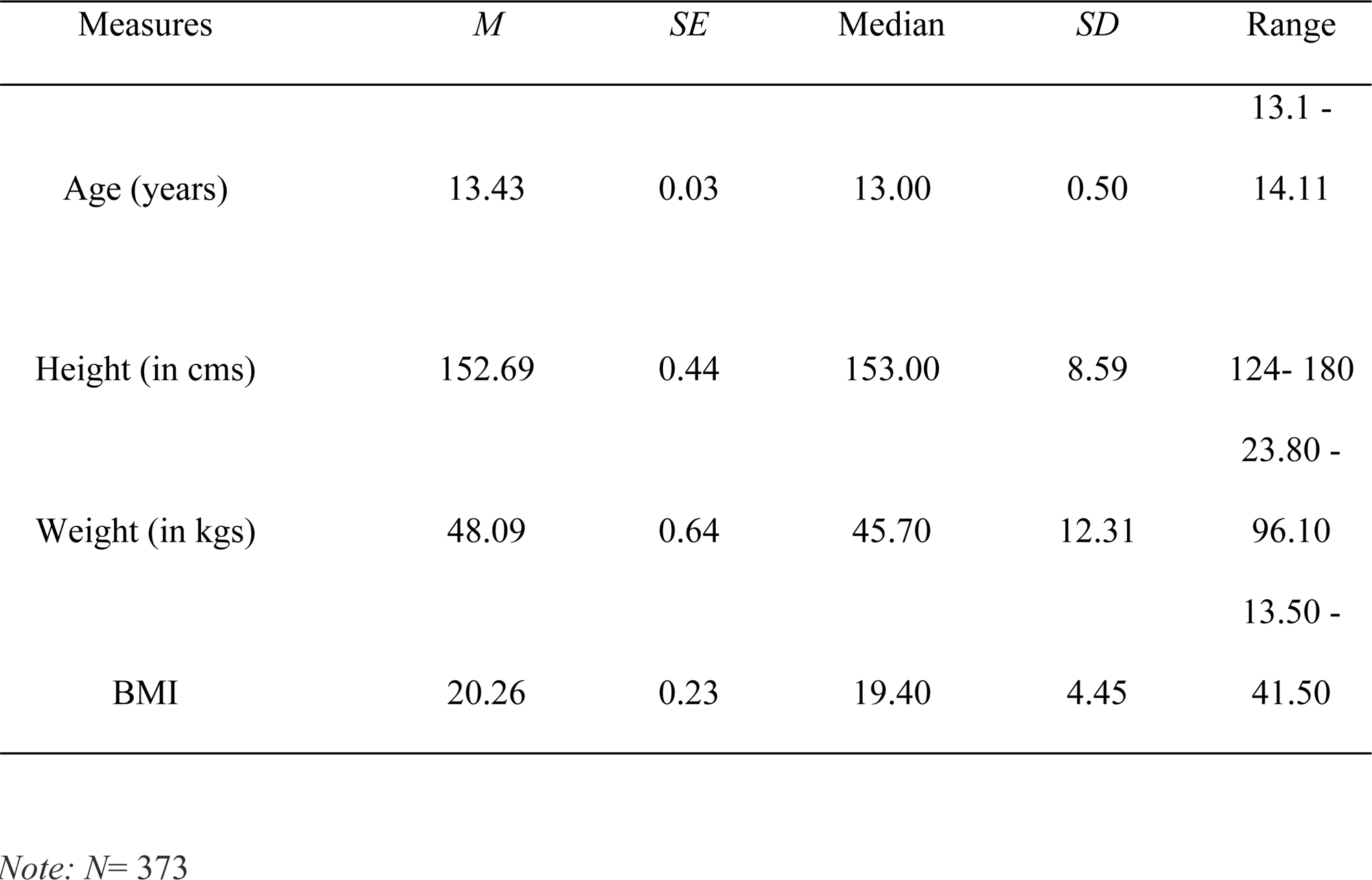
Socio-Demographic Factors.

### Body Mass Index

Based on the measurements, 39.7% (n= 149) of our participants were underweight, 46.1% (n= 173) had a normal BMI, 9.6% (n=36) were overweight, and 4.5% (n=17) were obese.

### Physical activity

All 373 participants were involved in some form of daily physical activity. 41.01% (n=153) performed aerobic exercises, 35.12% (n= 131) performed aerobics combined with balancing, and 13.3 % (n= 50) performed aerobic activities with resistance workouts. An exceedingly small percentage of children were involved in only balancing or only resistance workouts (5.6%, n=21), while only 1.9% (n = 7) performed all three types of activities. Moreover, 55% (n= 205) exercise for more than 30 minutes, 23% (n=86) exercise for 30 mins, and 21.9% (n= 82) exercise for less than 30 minutes daily. Note that playing outdoors was considered an aerobic activity.

### Sleeping Patterns of the Subjects

We found that 42.9% (n=161) of the participants usually slept for 8 hours a day, 41.2% (n=154) slept for less than 8 hours and 15.5% (n= 58) slept for more than 8 hours. It was also observed that 55.7% (n= 209) do not sleep during the day, while 44.3% (n= 166) of the participants usually sleep during the daytime. Of the total number of participants who sleep during the daytime, most of them take a nap for more than 30 minutes and a smaller percentage sleep for 30 minutes or less.

### Variety and Servings of Energy-Dense Foods

We found that the mean VEDF of the population was 27.98/week (SD: 16.33, median: 25), suggesting that these adolescents consumed approximately 4 varieties of EDF every day. Furthermore, they reported consuming a mean of 74.22 (SD: 48.4, median: 61) servings a week, indicating that they consumed approximately 10.60 servings of EDF a day. The mean variety per week of each food group is presented in Table 2:

**Table 2:** Mean variety/week of each food group.

| Food Groups | *Mean* | *SD* | *Median* |
| --- | --- | --- | --- |
| Chats and street foods | 7.69 | 4.52 | 7.0 |
| Main course | 3.12 | 12.11 | 2.0 |
| Starters | 2.52 | 2.31 | 2.0 |
| Aerated drinks and juices | 4.08 | 3.47 | 3.0 |
| Continental foods | 1.74 | 1.95 | 1.0 |
| Desserts and baked foods | 4.02 | 2.83 | 4.0 |
| Indian sweets | 2.58 | 2.74 | 2.0 |
| Farsan | 0.91 | 1.13 | 1.0 |
| Packaged snack items | 1.75 | 1.80 | 1.0 |

### Association between VEDF and zBMI

Regression analysis of our data showed no significant association (F-statistic: 2.262, p = 0.1334) between VEDF and BMI (Table3), indicating that consumption of VEDF was not a significant predictor of BMI in this group (**Fig1)**. Furthermore, after adding covariates and using multiple linear regression, the relationship between VEDF and BMI did not change. Since BMI is an ordinal variable, we also gauged linear by linear association analysis using Somer’s delta as a measure of agreement between predictors and the response variable, BMI for each of the food categories. It was observed that the variety and number of servings of all nine food groups were not associated with BMI.

**Fig1:**
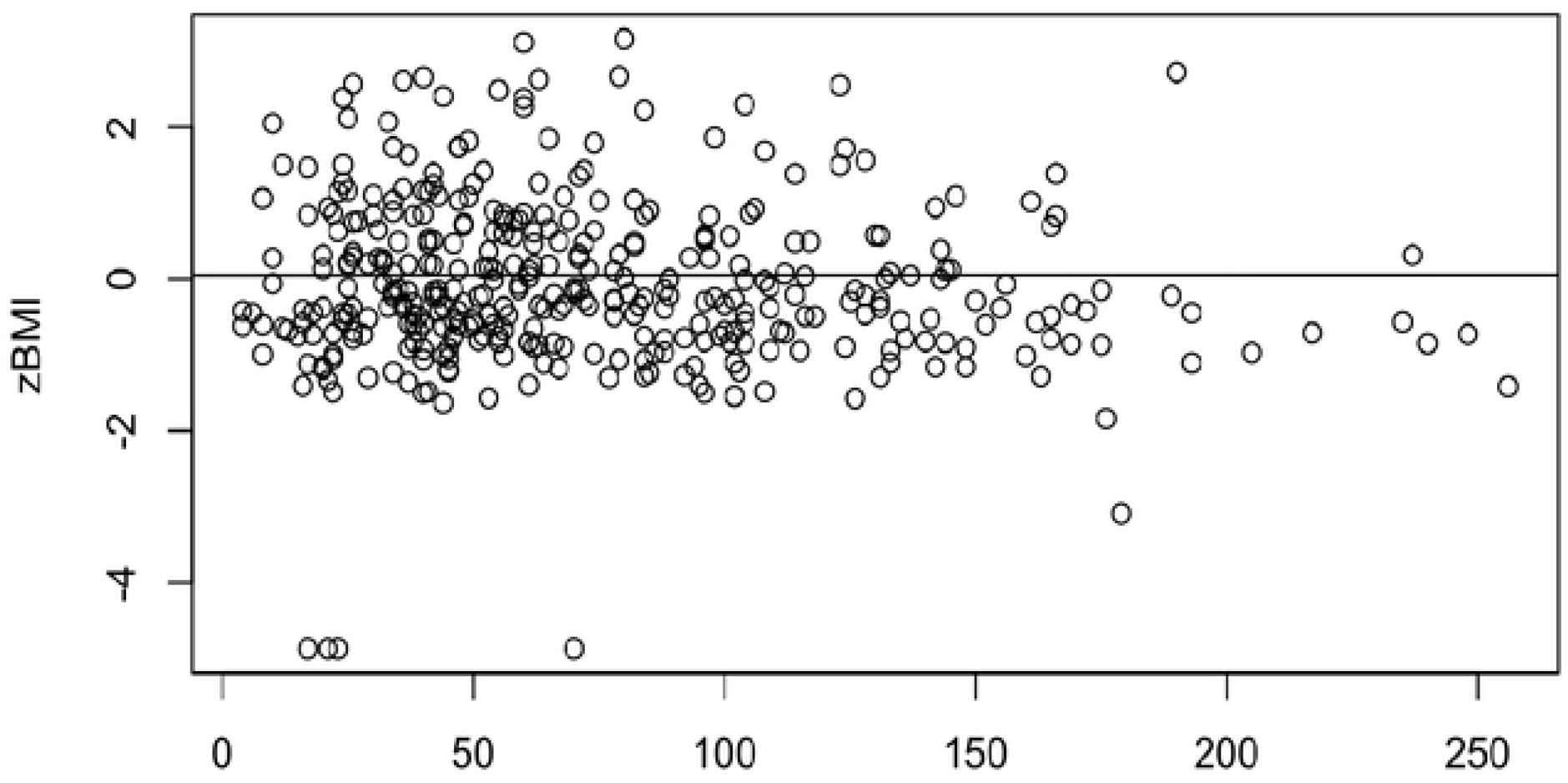
Relationship between zBMI and the variety of energy-dense foods consumed by 373 adolescents aged 13-15 years.

### Association between the number of servings of EDF and BMI

Linear regression of the number of servings revealed no significant association with BMI (F-statistic: 3.168, p=0.07592) (Table3), indicating no role of servings of EDF on BMI (Fig 2). To investigate the effect of VEDF on the total number of servings, regression analysis was also conducted between the two, and a positive significant association was detected (F-statistic: 2858, p: <2e-16). Based on our coefficient of determination (R2), we can state that around 88.43% of the change in the number of servings is accounted for by the VEDF. To verify, the variance inflation test using both VEDF and number of servings as predictors and BMI as the response variable was conducted, and it gave us results close to 1, indicating no multicollinearity between the two predictor variables.

**Fig 2:**
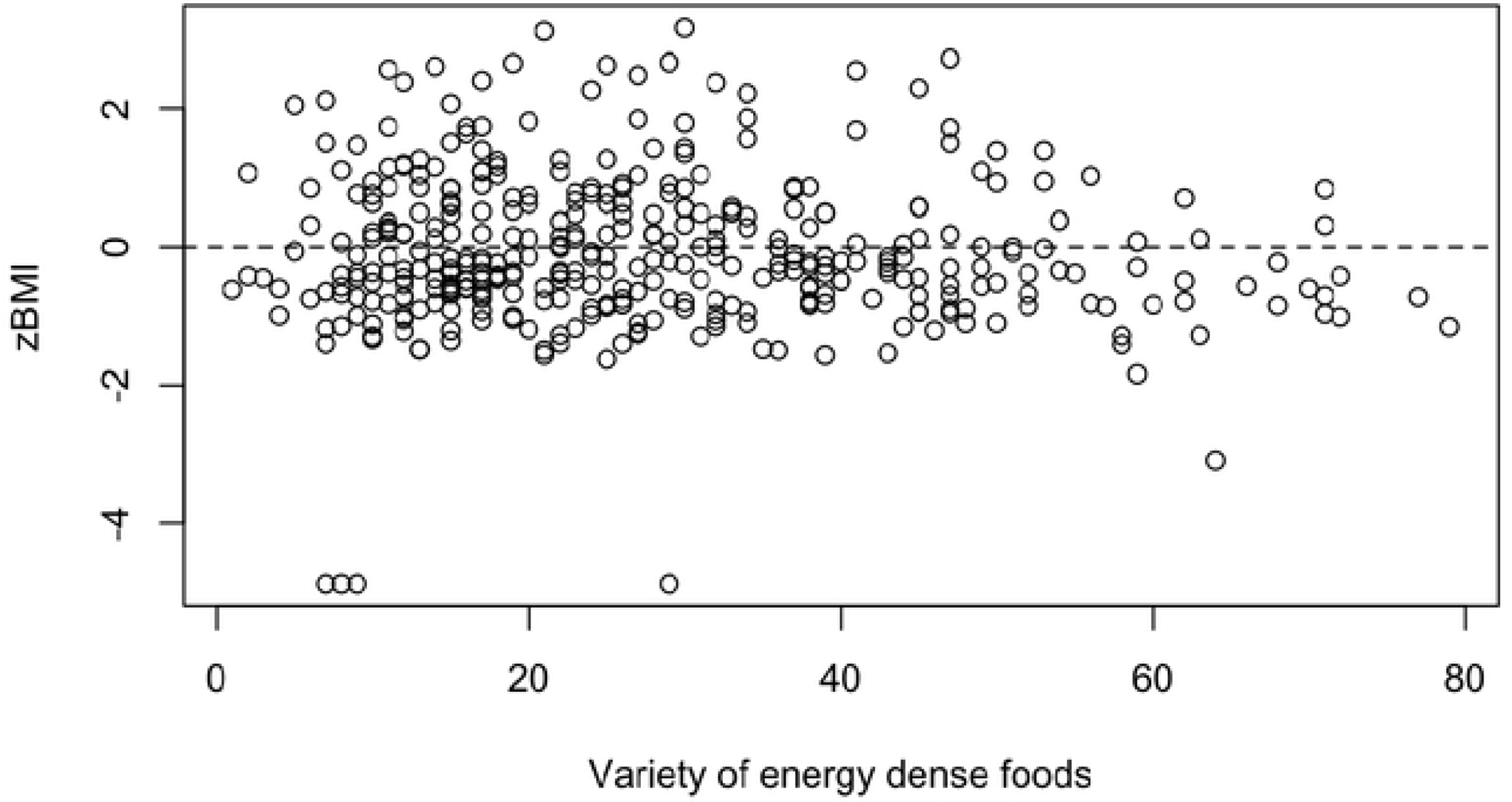
Relationship between zBMI and the number of servings of energy-dense foods consumed by 373 adolescents aged 13-15 years.

We further conducted the odds ratio for the number of servings of EDF to understand how it affects the odds of an increase in BMI. On analysis, we found the odds ratio of 0.99 indicating that there is no association between the occurrence of OW/O and the number of servings of EDF consumed in this population.

### Association between sleep and physical activity with BMI

Although spearman’s correlation found a significant inverse association between duration of sleep and zBMI (S= 9572, p-value= 0.0163, rho= −0.125), regression analysis revealed that there is no causative effect of sleep duration and zBMI in the present sample (F-value=2.402, p= 0.122) (Table3). Similarly, no significant association was found between the duration of exercise and BMI (F-value = 2.297, p= 0.1305) (Supplementary material). The number of times exercises were performed and their effect on BMI was also studied, and it was observed that the two are not significantly correlated. (F value= 2.019, p= 0.1562)

## Discussions

Given the rise in the prevalence of obesity among children and adolescents, effective preventative programs are needed to focus on the etiologic components that are most receptive to modification. In this study, we sought to examine the effect of one such component, that is, dietary variety of energy-dense foods, on BMI among adolescents living in Mumbai, India. On analysis, three key findings emerged from this investigation. First, no significant association was detected between VEDF and zBMI. Though there aren’t many research papers that have looked into the variety of EDF, our findings are consistent with several studies that found no correlation between the dietary energy density (DED) of foods and BMI in adolescents (32–36). The possible causes for the absence of association in this demographic are their high physical activity, which has been established to cancel the effect of EDF on their weight (37), an alarming number of underweight participants in the study, and the possibility of these participants compromising on regular, healthy meals by consuming EDF. It is imperative to also emphasize that the data for adults are far more consistent than for adolescents because a vast number of studies on adults have yielded clear conclusions (38,39). This implies that the processes by which DED occurs in adolescence differ from those reported both during adulthood when the predisposition for fat storage is larger (40), and in childhood when the response to fluctuations in energy density appears to occur innately (41,42).

The second key finding of the study is that participants consumed four varieties and ten servings of EDF on average per day, which is higher than the consumption reported in other countries (34,42). However, similar to our findings, a recent study conducted on adolescents residing in Kolkata, India, reported an intake of 6.25 servings of energy-dense snacks and 3 or more energy-dense beverages per day (42). Together, these two studies demonstrate that adolescents in India have poor dietary patterns. The fact that despite high consumption of EDF, the BMI of these adolescents is unaffected is especially a matter of concern due to the prevalence of the thin-fat phenotype in South Asian countries, including India. This phenotype causes individuals to have normal body weight (as measured by BMI) but a disproportionately high body fat percentage (44). There is evidence of a high risk associated with the epidemic of noncommunicable chronic diseases, and studies have also indicated that mortality associated with this phenotype is also significantly higher than in non-obese subjects (45). Additionally, it is known that Asians tend to have low BMI but high body fat percentage as compared to other ethnicities, which we hypothesize is the case with this population (46).

Third, no detectable association was found between the number of servings and adolescents’ zBMI. Though surprising, this finding is in line with studies in the past that have established that there is no “direct causal” link between portion sizes and obesity in the long term because energy intakes are a function of not only the portion size of food but also its energy density and the frequency of food and beverage consumption, among several other factors (47,48). However, we did find a strong positive association between VEDF and serving sizes, suggesting that consuming a variety of energy-dense foods augments the number of servings consumed by adolescents in this sample.

Our participants displayed some surprising characteristics. According to WHO, the global prevalence of obesity during adolescence in 2016 was 18% (49). Relative to those numbers, the prevalence of obesity in our study (4.5%) was much less. In fact, in the present study, many adolescents were underweight (39.7%) which is a matter of concern, especially given that these adolescents belonged to a higher socioeconomic status. The low prevalence of OW/O could also be attributed to the fact that all subjects regularly participate in aerobic physical activity, which is incorporated into the school schedule. This also explains the lack of correlation between VEDF and the number of servings of EDF with BMI as seen in the present study, since very few participants belonged to the OW/O category.

To our knowledge, the current study is the only one thus far that has attempted to investigate the relationship between VEDF, in specific, with BMI in Indian adolescents. Strengths of our study include a large sample size (n=373) and a high response rate. Given that adolescents frequently consume high-calorie foods and the fact that obesity is on the rise, the present study was also of public health relevance. We constructed indices of dietary variety to differentiate foods into categories that are commonly consumed by adolescents living in the given demographic.

Our analysis also has some important limitations. (i) The cross-sectional design of our study does not allow for causality conclusions, (ii) the self-reporting of information regarding dietary variety, portion sizes, sleep schedule, and physical activity is a source of bias. Moreover, since the foods are not weighed or quantified using household utensils in typical FFQs (Food Frequency Questionnaire), they tend to be less accurate compared to other quantitative dietary assessment methods including 24-hour Dietary Recalls and Weighed Food Records (50,51), (iii) We also had to curate our own FFQ because there are no questionnaires that focus only on VEDF. However, since the questionnaire is not a validated one, it adds to the limitations of our study. (iv) Additionally, adolescence is a time of critical reproductive and overall growth. Therefore, BMI alone isn’t sufficient, and measurement of body fat percentage would have served as a better indicator of OW/O.

Nonetheless, our results confirm the findings of previous, similar studies. Longitudinal data are necessary to further unravel the complex interplay between the outcome and the above-mentioned covariates. Adolescents could be randomized into distinct groups and supplemented with energy-dense foods in different varieties to see the causal relationship between VEDF and BMI. Additionally, future studies could assess the effects of VEDF on body fat percentage to elucidate the implications of excess EDF consumption and identify the presence of thin fat phenotype among Indian adolescents.

## Conclusion

The findings of the current study suggest that there is no significant association between the consumption of a variety of energy-dense foods and BMI, suggesting that VEDF does not play a key role in explaining the variability in BMI in this population. High levels of physical activity, a large percentage of underweight individuals in this demographic, and the difference in mechanisms by which adiposity affects adolescents could all contribute to this lack of association. This lack of relationship is consistent with previous studies that have investigated the effect of overall dietary energy density on BMI in adolescents. However, we did find that VEDF increases the number of servings consumed. This is a matter of concern since in urban areas, and with an open economy, the variety of food has increased and is readily available. Therefore, adolescents belonging to families with no financial constraints are particularly vulnerable. To conclude, longitudinal studies are warranted to see the impact of variety on BMI, and more so, on body composition.

## Data Availability

DOI: 10.5061/dryad.bg79cnpf8

## Acknowledgments

The authors express their gratitude to Anali Morales and Qianyue Wang from Purdue University for their valuable contributions in conducting statistical analysis of the data.

